# Remdesivir use in patients with coronavirus COVID-19 disease: a systematic review and meta-analysis of the Chinese Lancet trial with the NIH trial

**DOI:** 10.1101/2020.05.23.20110932

**Authors:** Paul Elias Alexander, Joshua Piticaru, Kimberley Lewis, Komal Aryal, Priya Thomas, Wojciech Szczeklik, Jakub Fronczek, Kamil Polok, Waleed Alhazzani, Manoj Mammen

## Abstract

**Background:** Coronavirus disease 2019 (COVID-19), caused by the novel coronavirus SARS-CoV-2, has led to significant global mortality and morbidity. Until now, no treatment has proven to be effective in COVID-19. To explore whether the use of remdesivir, initially an experimental broad-spectrum antiviral, is effective in the treatment of hospitalized patients with COVID-19, we conducted a systematic review and meta-analysis of randomized, placebo-controlled trials investigating its use.

**Methods:** A rapid search of the MEDLINE and EMBASE medical databases was conducted for randomized controlled trials. A systematic approach was used to screen, abstract, and critically appraise the studies. Grading of Recommendations Assessment, Development, and Evaluation (GRADE) method was applied to rate the certainty and quality of the evidence reported per study.

**Results:** Two RCTs studies were identified (n=1,299). A fixed-effects meta-analysis revealed reductions in mortality (RR=0.69, 0.49 to 0.99), time to clinical improvement (3.95 less days, from 3.86 days less to 4.05 less days), serious adverse events (RR=0.77, 0.63 to 0.94) and all adverse events (RR=0.87, 0.79 to 0.96).

**Conclusion:** In this rapid systematic review, we present pooled evidence from the 2 included RCT studies that reveal that remdesivir has a modest yet significant reduction in mortality and significantly improves the time to recovery, as well as significantly reduced risk in adverse events and serious adverse events. It is more than likely that as an antiviral, remdesivir is not sufficient on its own and may be suitable in combination with other antivirals or treatments such as convalescent plasma. Research is ongoing to clarify and contextual these promising findings.

## Background

Previous human coronaviruses, which resulted in high-morbidity (and mortality), such as the acute respiratory syndrome coronavirus (SARS-CoV or SARS-1) in 2003 and the Middle East respiratory syndrome coronavirus (MERS-CoV) in 2012, have driven the development of compounds that could potentially be active against the coronavirus disease 2019 (COVID-19) (caused by the novel coronavirus SARS-CoV-2).^1-4^

Approximately 15% of the patients with confirmed COVID-19 exhibit severe symptoms and require hospitalization, and many of them requiring hospitalization may need intensive care resources.^5^ The current number of cases worldwide is over five million, with over 275,000 deaths reported as of May 2020.^6^ Case fatality (mortality) rates reported to date based on laboratory confirmed cases are likely to overestimate the actual COVID-19 mortality rate as they did not include mild or asymptomatic cases that have recovered. Ideally, the infection fatality rate should be reported as comprehensive data is available which would be important to inform planning and response.

Currently, no treatment nor prophylaxis has proven to be effective in COVID-19, and patients receive either symptomatic treatment for milder presentations or more advanced life-support strategies in moderate to severe cases. These include oxygenation strategies in a hospitalized ICU setting. Severe disease results in a rapid progression of respiratory failure that begins usually right after dyspnea (breathing difficulties) and hypoxemia. A cytokine release storm with elevated blood inflammatory markers is a cardinal feature and progression to severe illness can result in severe organ failure with forms of kidney, liver, and cardiac pathology and injury and indications of coagulopathy and potential shock. As the global community eagerly awaits credible scientific solutions for this pandemic, researchers and scientists are working hard to identify effective therapeutic and preventive strategies for COVID-19.

Recent research findings on the possible therapeutics such as antivirals, antimalarials such as chloroquine or hydroxychloroquine, in COVID-19 patients have been mostly vague, inconclusive or negative, typically underpinned by very low-quality research methodologies and inadequate reporting.^7,8^ The very poor methodological quality thus far across the COVID-19 research landscape raises many concerns as the medical research community and governments urgently seek an effective prophylactic or therapeutic. The methodologies and reporting in these studies have been suboptimal. The randomized controlled trials (RCTs) have been plagued with sub-optimal randomization, a lack of allocation concealment, no or questionable blinding, small sample sizes, small event numbers, and sparse reporting. The observational studies have been confounded, had small sample sizes, small numbers of events, sub-optimal or absent statistical adjustments, matching (propensity score approach), stratification, or any methodological technique that could mitigate the risk of biased estimates of effect. Precious time and vast amount of resources are potentially being squandered as the trustworthy, robust, high-quality research that is needed for evidence-informed decision-making and policy decisions is side-lined.

This urgency of clinical research has included the repurposing of existing therapies for other diseases based on indirect evidence. For example, the use of corticosteroids in COVID-19 is based on the therapeutic benefit seen in randomized controlled trials (RCTs) of corticosteroid use in patients with acute respiratory distress syndrome (ARDS)^9-11^.

Initial evidence on the apparent efficacy of remdesivir against Middle Eastern Respiratory Syndrome (MERS) and SARS-CoV-2 from in-vitro models helped remdesivir gain attention as a possible therapeutic agent.^12,13^ Prior to the pandemic, remdesivir was an unapproved investigational product. A recent observational study examining the compassionate use of remdesivir in patients with severe COVID-19 revealed that 68% of patients had clinical improvement. ^14^ In April, the first RCT of remdesivir in patients with severe COVID-19 was published; however, it was not able to identify clinical benefit and had a small sample size and failed to reach the enrolment target.

In early May 2020, the US Federal Drug Administration (FDA) authorized the use of remdesivir for the treatment of individuals with severe COVID-19 based on preliminary data from the ACTT-1 study. Due to the rapid spread of COVID-19 with its concerning morbidity and mortality, it is unlikely we will see further randomized, placebo-controlled, double-blind trials in light of existing data. Given the urgency for an effective therapeutic, we conducted a rapid review of the existing medical research literature assessing studies using remdesivir in RCTs in patients with COVID-19.^15,16^ We were alert to and considered all studies published since the global pandemic began in January 2020, while recognizing that the two RCTs would likely comprise our final cohort of included studies.

## Methods

### Search strategy and study selection

MEDLINE and EMBASE electronic databases were searched from January 1st, 2020, to May 22nd, 2020, using a mix of keywords such as ‘COVID-19’ and ‘remdesivir along with any relevant variants. For example, the COVID-19 search terms considered were ‘exp Coronavirus Infections/ or exp Coronavirus/ or exp Severe Acute Respiratory Syndrome/ or exp SARS Virus/ or coronavirus.mp. or severe acute respiratory syndrome coronavirus 2.mp. or 2019 nCoV.mp. or 2019nCoV.mp. or 2019 novel coronavirus.mp. or new coronavirus.mp. or novel coronavirus.mp. or SARS-CoV-2.mp. or SARS CoV-2.mp. or COVID 19.mp. or COVID-19.mp. or COVID19.mp.’ The search did use an RCT filter (see Appendix for MEDLINE search example). PubMed was also searched daily during this period to rapidly assess any emerging publications. Evidence was considered from additional sources, including Google Scholar and online preprint servers, that pre-publish studies not yet having completed the peer-review process. We also searched the largest clinical medicine preprint repository, medRxiv.org.

### Data Abstraction and Quality Assessment

Two reviewers independently and in duplicate, extracted relevant data, and assessed risk of bias using the Cochrane risk of bias tool for RCTs.^17^ We adopted response options for risk of bias to be ‘yes’, ‘probably yes’, ‘probably no’, and ‘no’, thereby removing the ‘unclear’ response often reported by reviewers. The use of the ‘unclear’ response frequently hampers a more definitive interpretation of risk of bias. ^18^

### Data Synthesis and Analysis

We used fixed-effect (inverse-variance) modeling for all analyses (Mantel–Haenszel risk ratio (RR) for dichotomous outcomes and mean difference (MD) for continuous variables).^19^ Our intended modelling decision made *a priori* was based on the potential small number of RCTs we would deem eligible and thus the decision to model whereby more weight would be given to larger sample sized studies. We would however revert to a random-effects approach should the number of uncovered RCTs be high (though we did not anticipate this). When data were reported as medians and inter-quartile ranges, we would convert them to means and standard deviations for meta-analytical pooling.^20^ We also planned to conduct sensitivity analyses based on random and fixed effect modeling.

Review Manager 5.3 was used to conduct the meta-analysis.^21^ We assessed heterogeneity by visual inspection of forest plots, a Cochrane chi-square statistical test for heterogeneity, and the *I*_2_ statistic (with >50% considered as significant heterogeneity warranting exploration and explaining).^22^ We report 95% confidence interval (CI) measures of uncertainty with presented estimates of effect. To estimate the baseline risk in computing the absolute effects, we used the control event rate. To compute the absolute effect, we multiplied the baseline risk by the relative effect (and 95% CIs).

### GRADE Methods

We utilized the *Grading of Recommendations Assessment, Development, and Evaluation* (GRADE) approach to rate the certainty of the evidence for each outcome and the entire body of evidence. ^23^ This outcome centric approach for rating effect estimate certainty considers the body of evidence via risk of bias, imprecision, inconsistency (heterogeneity), indirectness, and publication bias domains when assessing RCTs.

## Results

### Study characteristics

The search uncovered two RCT citations, and with rapid screening and review, those two were deemed eligible to inform the final review (Table 1).^15,16^ One was a multicenter study conducted in China, and the second study was an international, multicenter trial lead by the National Institutes of Health (NIAID/NIH) of the USA. The ages ranged from 58 years old to 66 years old, and the percentage of males ranged from 56% to 65%. There were 135 deaths out of a total of 1,299 enrolled patients.

**Table 1:** Characteristics of included studies.

| Author, year, location, study type | Intervention, Age, Gender | Intervention, sample size | Intervention, Time from symptom onset to starting randomization/study treatment, days | Intervention, Additional Therapy (%) | Intervention, Time to clinical improvement | Intervention, Mortality (%) | Intervention, Serious adverse events (%) | Risk of bias assessment |
| --- | --- | --- | --- | --- | --- | --- | --- | --- |
|  | Placebo, Age, Gender | Placebo, sample size | Placebo, Time from symptom onset to starting study treatment, days | Placebo, Additional Therapy (%) | Placebo, Time to clinical improvement | Control, Mortality (%) | Intervention, Serious adverse events (%) |  |
| Wang, 2020,<sup>15</sup> China, multicenter, randomized, double-blind, placebo-controlled | Median 66 (IQR 57-73)<br>Male 56% | Remdesivir, 158 | Median 11 (IQR 9-12) | Interferon alfa-2b (29%), lopinavir-ritonavir (28%), corticosteroids therapy (65%) | Median 21·0 (IQR 13·0-28·0) | 28-day mortality Remdesivir, 22 (14%) | 28 (18%) | Low-moderate |
|  | Median 64 (IQR 53-70)<br>Male 65% | Placebo, 78 | Median 10 (IQR 9-12) | Interferon alfa-2b (38%), lopinavir-ritonavir (39%), corticosteroids therapy (68%) | Median 23 (IQR 15·0-28·0) | 28-day mortality Placebo, 10 (13%) | 20 (26%) |  |
| Biegel, 2020,<sup>16</sup> multinational, multicenter, randomized, double-blind, placebo-controlled | Mean 58·6 (SD 14·6)<br>Male 65% | Remdesivir, 541 | Median 9 (IQR 6-12) | Not reported | Median 11 (95% CI 9-12) | 14-day mortality Remdesivir, 32 (7·1%*) | 114 (21%) | Low-moderate |
|  | Mean 59·2 (SD 15·4)<br>Male 63% | Placebo, 522 | Median 9 (IQR 7-13) | Not reported | Median 15 (95% CI 13-19) | 14-day mortality Placebo, 54 (11·9%*) | 141 (27%) |  |

Abbreviations used: IQR- interquartile range, SD- standard deviation, Serious adverse events a detailed number of patients with serious adverse events
Footnote: \* Kaplan-Meier estimator was used to calculate the mortality rates due to patients still in follow-up before publication of preliminary results. Our decision was guided by the study authors reporting the Hazard Ratio that was non-significant for mortality, and this did not correspond to the raw mortality data in the outcomes table.

### Risk of bias assessment

Both RCTs were judged to be of a low-moderate risk of biased estimates ^15,16^ of effect, despite the study reported by Biegel having a change in the primary outcome before unblinding, and the loss of data reported due to publication of preliminary results before full follow-up was completed on all study participants. Otherwise, the methodologies were robust, and reporting was adequate. We also developed no funnel plots or statistical tests for publication bias due to the limited interpretability when the study number is less than the optimal threshold of 10.^24^

## Outcomes

### Mortality

The two studies reported on mortality, but the Wang study15 reported 28-day mortality, and the Biegel study^16^ only reported on 14-day mortality. Due to the Biegel study reporting preliminary results, with many subjects still within the follow-up phase of the trial, the mortality estimates were reported on the Kaplan-Meier estimator.

Based on our reading of the reported mortality findings by Biegel^16^ and the pooling of the 14-day mortality from the Biegel16 study with Kaplan-Meier estimates to the Wang15 mortality data (n=1,141 patients), there was a significant reduction of mortality in patients who received remdesivir compared to the control group (RR 0·69, 95% CI 0·49 to 0·99), *p*<0.05; *I*_2_ = 52%; fixed-effect modeling, low certainty) (Figure 1). Absolute effects were 35 fewer deaths per 1,000 affected individuals (54 fewer to 1 fewer) (GRADE evidence profile Table 2). Sensitivity analysis using random-effects modeling and the use of intention to treat (ITT) population for Biegel^16^, or the use of the sample size reported as available for analysis, reveals no difference in direction or magnitude of risk estimates. In fact, when the raw mortality data is utilized in fixed-effect modelling with the reported full sample size, we found a more significant reduction in mortality due to remdesivir (RR 0.67, 95% CI 0.47 to 0.96). However, as indicated, we modelled based on the Kaplan-Meier estimator.

**Table 2;.**
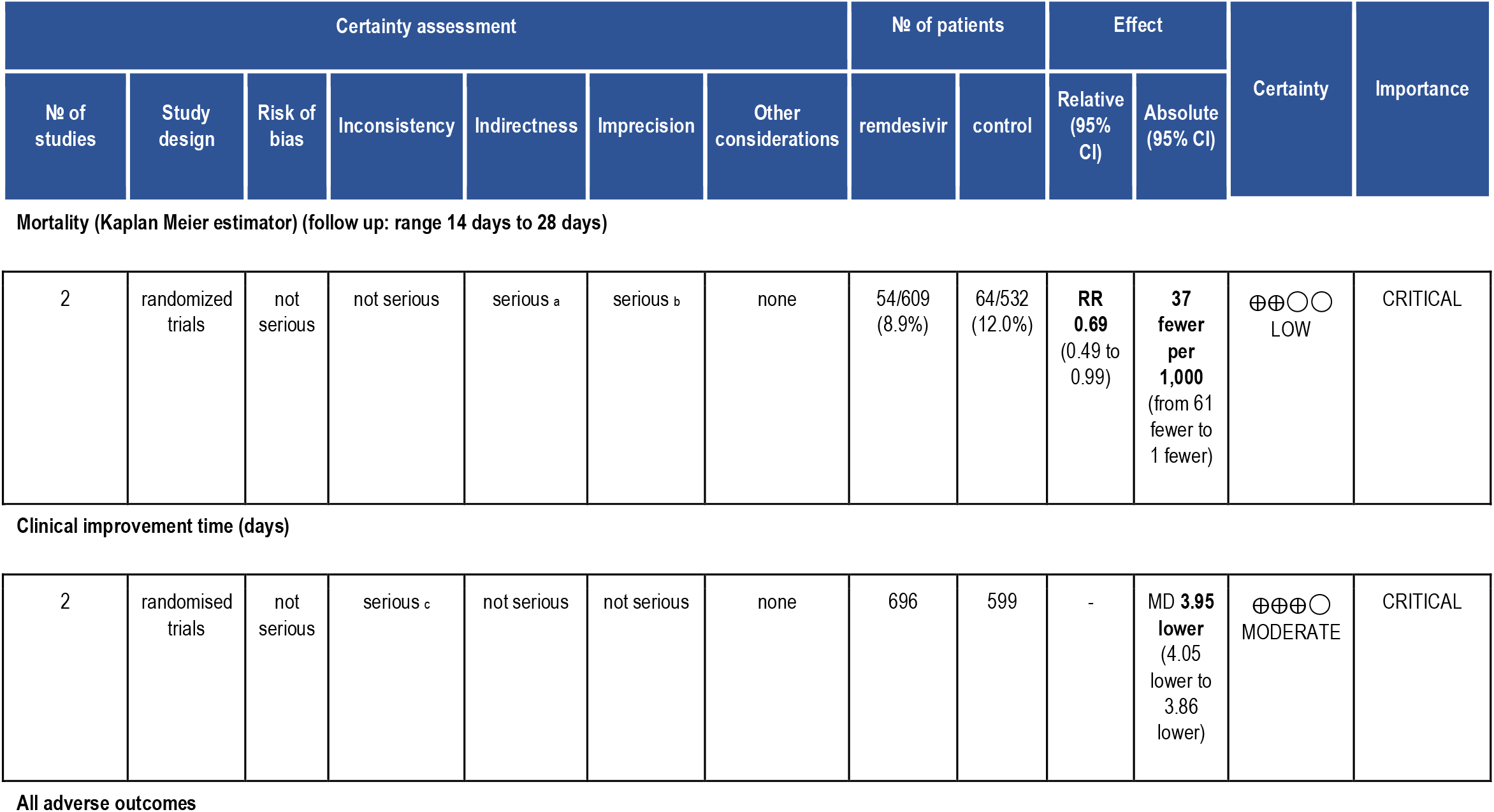
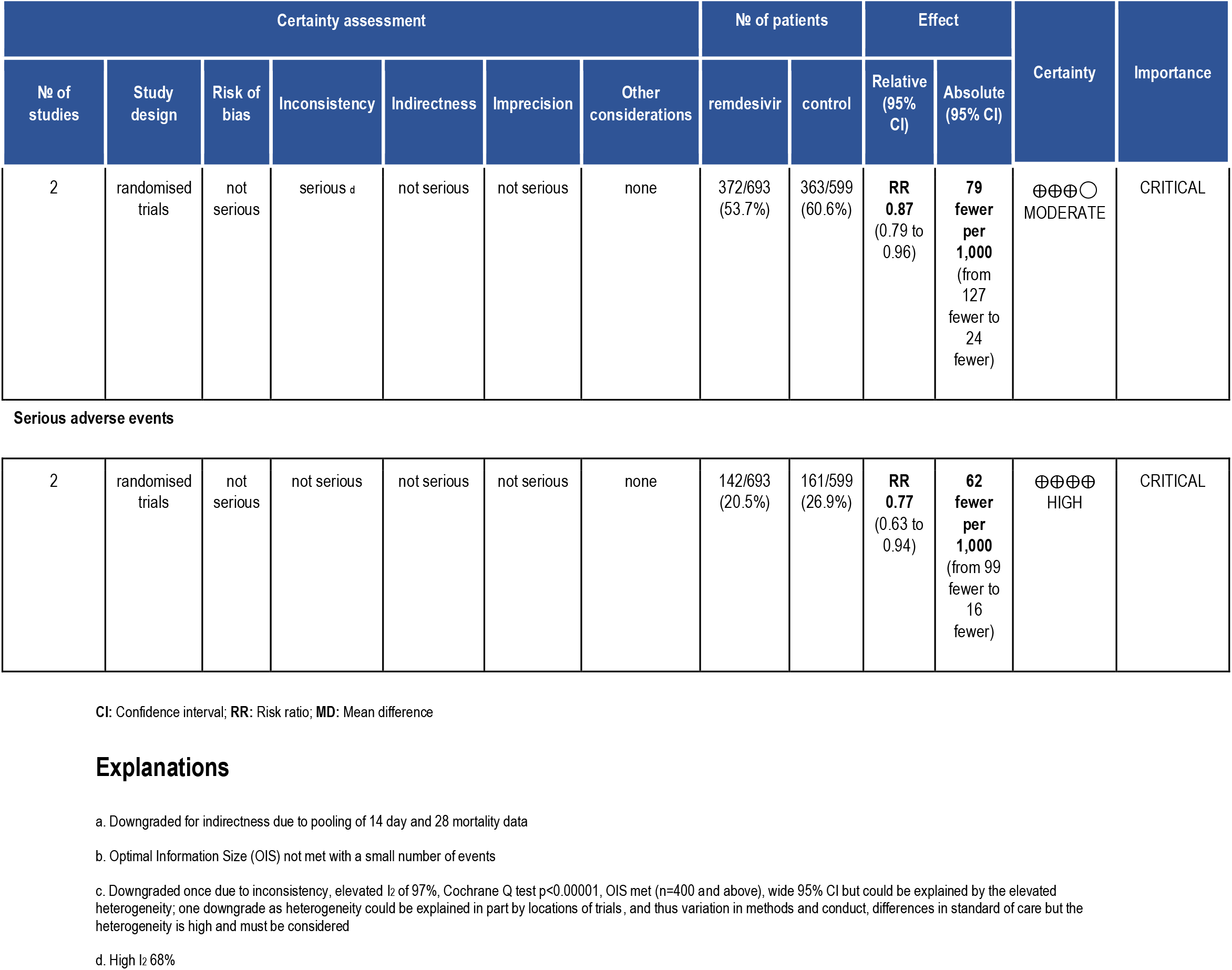
GRADE evidence table

| Certainty assessment |  |  |  |  |  |  | No of patients |  | Effect |  | Certainty | Importance |
| --- | --- | --- | --- | --- | --- | --- | --- | --- | --- | --- | --- | --- |
| No of studies | Study design | Risk of bias | Inconsistency | Indirectness | Imprecision | Other considerations | remdesivir | control | Relative (95% CI) | Absolute (95% CI) |  |  |
| **Mortality (Kaplan Meier estimator) (follow up: range 14 days to 28 days)** |  |  |  |  |  |  |  |  |  |  |  |  |
| 2 | randomized trials | not serious | not serious | serious <sup>a</sup> | serious <sup>b</sup> | none | 54/609 (8.9%) | 64/532 (12.0%) | RR 0.69 (0.49 to 0.99) | 37 fewer per 1,000 (from 61 fewer to 1 fewer) | ⊕⊕○○ LOW | CRITICAL |
| **Clinical improvement time (days)** |  |  |  |  |  |  |  |  |  |  |  |  |
| 2 | randomised trials | not serious | serious <sup>c</sup> | not serious | not serious | none | 696 | 599 | - | MD 3.95 lower (4.05 lower to 3.86 lower) | ⊕⊕⊕○ MODERATE | CRITICAL |

**All adverse outcomes**
| Certainty assessment |  |  |  |  |  |  | № of patients |  | Effect |  | Certainty | Importance |
| --- | --- | --- | --- | --- | --- | --- | --- | --- | --- | --- | --- | --- |
| № of studies | Study design | Risk of bias | Inconsistency | Indirectness | Imprecision | Other considerations | remdesivir | control | Relative (95% CI) | Absolute (95% CI) |  |  |
| 2 | randomised trials | not serious | serious <sup>d</sup> | not serious | not serious | none | 372/693 (53.7%) | 363/599 (60.6%) | RR 0.87 (0.79 to 0.96) | 79 fewer per 1,000 (from 127 fewer to 24 fewer) | ⊕⊕⊕○ MODERATE | CRITICAL |
| Serious adverse events |  |  |  |  |  |  |  |  |  |  |  |  |
| 2 | randomised trials | not serious | not serious | not serious | not serious | none | 142/693 (20.5%) | 161/599 (26.9%) | RR 0.77 (0.63 to 0.94) | 62 fewer per 1,000 (from 99 fewer to 16 fewer) | ⊕⊕⊕⊕ HIGH | CRITICAL |

CI: Confidence interval; RR: Risk ratio; MD: Mean difference

**Figure 1:**
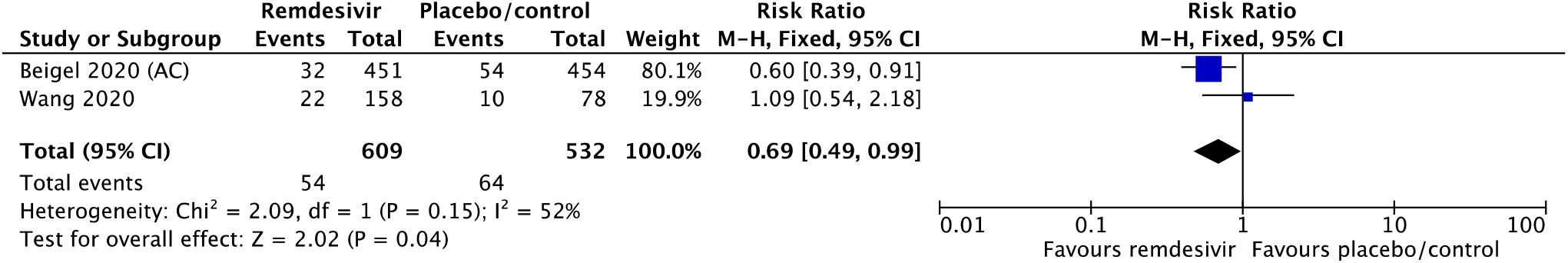
Mortality using remdesivir in hospitalized patients with COVID-19

**Figure 2:**
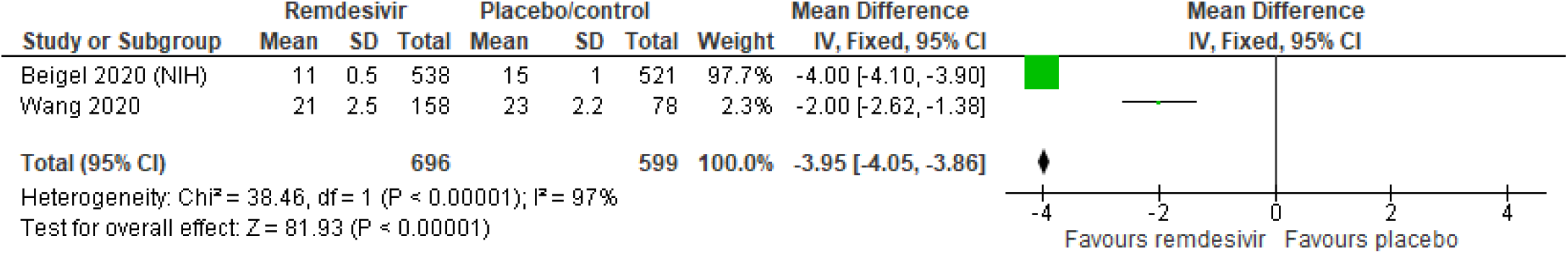
Time to clinical improvement using remdesivir in hospitalized patients with COVID-19

### Time to Clinical Improvement

Two studies (n=1,299) reported on time to clinical improvement and revealed significant improvement of time to clinical improvement in patients who received remdesivir compared to the control group (mean difference (MD) -3·95 days, 95% CI -4.05 days to -3.86 days), *p*=0.11; *I*_2_ = 97%; fixed-effect modeling, moderate certainty) (Figure 1). Absolute effects were 3·95 fewer days (4.05 fewer days to 3.86 fewer days) (GRADE evidence profile Table 2). Sensitivity analysis using random-effect modeling reveals a wider confidence interval but no difference in the direction of risk estimates.

### Serious Adverse Events

Two studies (n=1,299) reported on the number of patients with adverse events and revealed a significant reduction in serious adverse events in patients who received remdesivir compared to the control group (RR 0·77, 95% CI 0·63 to 0·94), *p*=0.01; *I*_2_ = 0%; fixed-effect modeling, moderate certainty) (Figure 3). Absolute effects were 62 fewer people with adverse events per 1,000 affected individuals (99 fewer to 16 fewer) (GRADE evidence profile Table 2). Sensitivity analysis using random-effects modeling reveals no difference in the direction of risk estimates.

**Figure 3:**
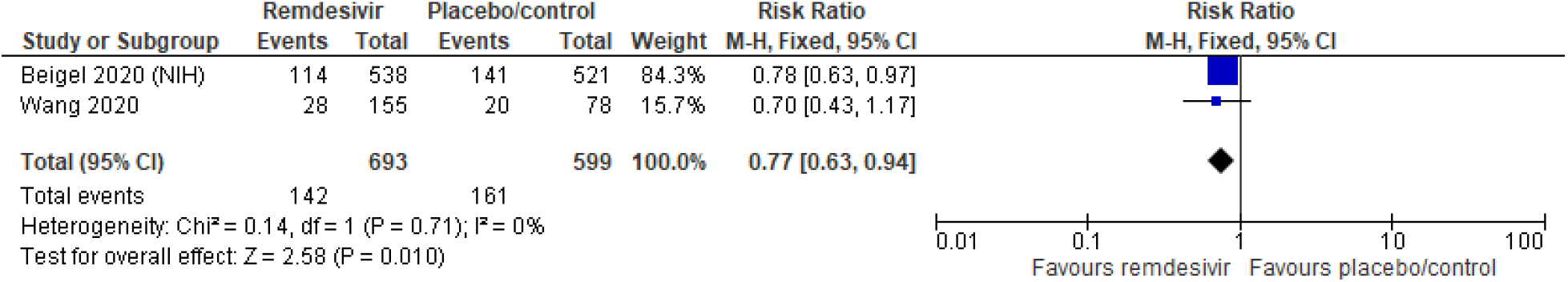
Number of patients with serious adverse events in hospitalized patients with COVID-19

### All Adverse Events

Two studies (n=1,299) reported on the number of patients with all adverse events and revealed a significant reduction in serious adverse events in patients who received remdesivir compared to the control group (RR 0·87, 95% CI 0·79 to 0·96), *p*<0.01; *I*_2_ = 68%; fixed-effects modeling, moderate certainty) (Figure 4A). Absolute effects were 79 fewer people with adverse events per 1,000 affected individuals (127 fewer to 24 fewer) (GRADE evidence profile Table 2). Sensitivity analysis using random-effects modeling reveals a wider confidence interval that crosses into harms (Figure 4B).

**Figure 4A:**
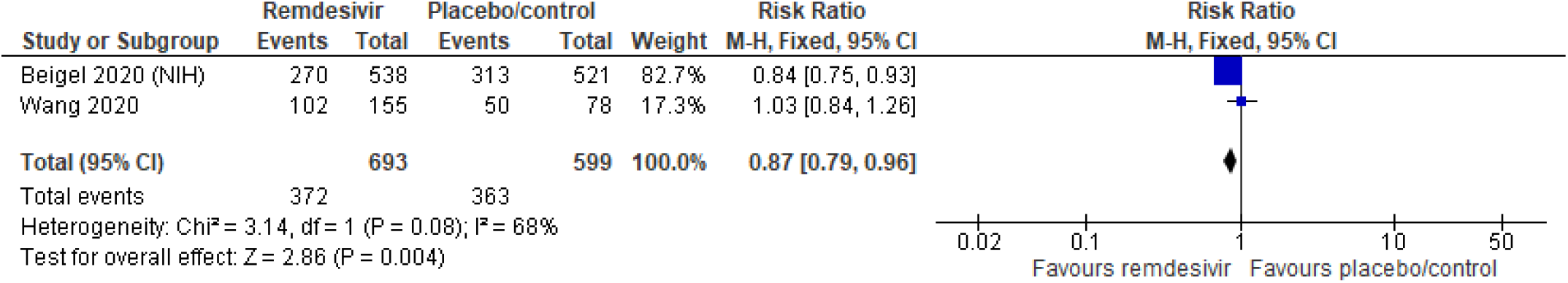
Number of patients with any adverse events in hospitalized patients with COVID-19 (fixed effects modeling)

**Figure 4B:**
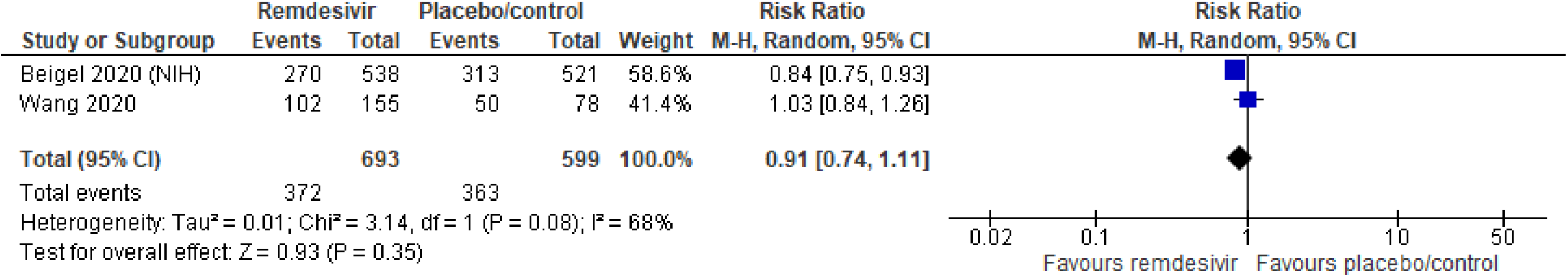
Number of patients with any adverse events in hospitalized patients with COVID-19 (random-effects modeling)

### Explanations

a. Downgraded for indirectness due to pooling of 14 day and 28 mortality data

b. Optimal Information Size (OIS) not met with a small number of events

c. Downgraded once due to inconsistency, elevated I2 of 97%, Cochrane Q test p<0.00001, OIS met (n=400 and above), wide 95% CI but could be explained by the elevated heterogeneity; one downgrade as heterogeneity could be explained in part by locations of trials, and thus variation in methods and conduct, differences in standard of care but the heterogeneity is high and must be considered

d. High I^2^ 68%

## Discussion

The two randomized-controlled trial studies presented in this systematic review focused on remdesivir use in hospitalized COVID-19 patients. Our pooled analysis reveals that remdesivir significantly reduces mortality, time to clinical improvement, the number of people with serious adverse events, and the number of people with any adverse events.

The GRADE quality of evidence (also known as certainty or confidence in estimates of effect) reflects the extent of our certainty or confidence that the estimates of an effect are adequate to support a particular decision or recommendation.23 We found some issues applying GRADE with indirectness, imprecision, and inconsistency across the body of evidence for the analyzed patient-important outcomes. We have low certainty or confidence in the range of mortality effects despite reportedly significant benefits of a decrease in mortality, with 37 fewer deaths per 1,000, with a range from 61 fewer to 1 fewer death; the range of mortality absolute risk effect ranges from a 51% reduction to a 1% reduction of risk. The low certainty in the risk of mortality estimates are due to the serious concerns with imprecision due to a small number of events, and indirectness due to the pooling of 14 day and 28-day mortality results. We made the judgement to pool mortality data based on the hazard ratio Kaplan-Meier data and not simply the raw data reported in Table 2 (32 in remdesivir vs 54 in placebo with samples sizes of 538 and 521 respectively; incidentally, mortality reductions would have been even more favourable). We employed a fix-effect model given the small number of studies and a decision to focus more weight on the study with the larger sample size (contributes more information).

We are moderately confident remdesivir in patients hospitalized with COVID-19 leads to faster clinical improvement, with an important and significant decrease in time to clinical improvement using remdesivir of 3.95 fewer days, ranging from 4.05 fewer to 3.86 fewer days. We are highly confident in an important and significant decrease in severe adverse events with the use of remdesivir in the individuals hospitalized with COVID-19, with 62 fewer severe events per 1,000 ranging from 99 fewer to 16 fewer serious adverse events.

Using random effect modelling did not change the pooled point estimates or confidence intervals appreciably in the context of harms and benefits of the intervention for all reported outcomes, except for all adverse events. For all adverse events, the use of random-effects modeling led to a confidence interval that spanned harms and benefits; with fixed-effect modeling, the confidence interval was wholly in the benefits portion, leading us to be moderately certain in a modest reduction in adverse events. Given the reduction in imprecision for all adverse events outcome, the GRADE assessment was moderate with fixed effect modeling and low with random-effects modeling.

Our study has several strengths. Firstly, we assessed the eligibility of all studies and the risk of bias independently and in duplicate. We intended the summary pooling of the data had we judged that the data was reliable enough to allow a meaningful pooled estimate of effect. Additionally, we intended the use of the GRADE approach to assess the certainty or quality of the evidence. Our analysis of the available literature had significant limitations, including the paucity of studies available for analysis. The research evidence on the benefits of remdesivir in COVID-19 is however, in our judgement, based on two rigorous multicenter trials.

Additionally, there was significant heterogeneity in the time to clinical improvement, which could be due to the location of the studies and the differences in standard of care between the studies that were conducted at different periods in the context of the evolving evidence landscape during the pandemic. Timing of initiation of remdesivir may also impact clinical outcomes, with patients with more severe disease being less likely to survive as seen in the ACTT-1 subgroups, however we were unable to pool data according to these subgroups.

Due to the public health emergency of the COVID-19 pandemic, there likely will not be any additional randomized, placebo-controlled, double-blind trial to determine remdesivir’s efficacy in patients with COVID-19, and instead, it would be incorporated into standard of care. In addition, remdesivir appears to reduce the time to clinical improvement and the number of serious adverse events and can ameliorate the conditions at many overwhelmed hospitals facing the pandemic patient surges.

The primary concern present in the context of the current crisis with continued lack of a viable treatment option in patients suffering from Sars-CoV-2 life-threatening respiratory failure, indeed persists. We are still left with questions, and clinicians must interpret this pooled evidence with caution, though very promising. Further research on the use of other therapeutic agents in COVID-19 patients is urgently needed and it is more likely that remdesivir would be of most utility when used in combination with other potential therapeutics (based on hopefully positive findings from ongoing studies). Such robust studies should be adequately powered RCTs, with longer durations of follow-up, that are methodologically superior, and focuses on harm outcomes.

## Conclusion

The primary concern present in the context of the current crisis with continued lack of a viable treatment option in patients suffering from Sars-CoV-2 life-threatening respiratory failure, indeed persists. We are still left with questions, and clinicians must interpret this pooled evidence with caution, though these data are very promising. Further research on the use of other therapeutic agents in COVID-19 patients is urgently needed and it is more likely that remdesivir would be of most utility when used in combination with other potential therapeutics (based on hopefully positive findings from ongoing studies), or perhaps when used earlier in the disease course. Such robust studies should be adequately powered RCTs with longer durations of follow-up, sound methodology and with focuses on harm outcomes.

In closing, we remind that safety of the patient suffering from COVID19 is a key priority to improve the quality of care in the provision of health services. Currently, no therapeutic option has been shown to be effective (though remdesivir is presently revealing promise as one option likely in conjunction with another treatment (s)) and that conclusively allows for safe and effective use to mitigate or eliminate the causative agent of COVID-19; The very same can be said about prophylaxis whereby approximately 200 therapeutic options or their combinations are being investigated in more than 1,700 clinical trials. Severely ill patients with COVID-19, frequently older adults and with established comorbidities, are being given multiple concomitant medications and without considering possible adverse events and interactions. This is an area of research that is being overlooked and the potential toxicity due to concomitant treatments must be urgently addressed. We again remind that the use of medications such as chloroquine, hydroxychloroquine (alone or in combination with azithromycin), ivermectin, antivirals, and immunomodulators, among others, should be considered within the context of patient consented, randomized clinical trials that evaluate their safety and efficacy. Ensuring the safety of patients with COVID-19 requires information and surveillance systems that include standardized procedures in order to report adverse events and interactions according to local settings and public health reporting regulations. If these do not exist, they must be implemented immediately.

## Data Availability

All data used for this systematic review is publically available.No F

